# Improving Upper-limb Prosthesis Usability: Cognitive Workload Measures Quantify Task Difficulty

**DOI:** 10.1101/2022.08.02.22278038

**Authors:** Michael D. Paskett, Jhorg K. Garcia, Sonny T. Jones, Mark R. Brinton, Tyler S. Davis, Christopher C. Duncan, Joel M. Cooper, David L. Strayer, Gregory A. Clark

## Abstract

Providing user-focused, objective, and quantified metrics for prosthesis usability may help reduce the high (up to 50%) abandonment rates and accelerate the clinical adoption and cost reimbursement for new and improved prosthetic systems. We comparatively evaluated several physiological, behavioral, and subjective cognitive workload measures applied to upper-limb neuroprosthesis use.

Users controlled a virtual prosthetic arm via surface electromyography (sEMG) and completed a virtual target control task at easy and hard levels of difficulty (with large and small targets, respectively). As indices of cognitive workload, we took behavioral (Detection Response Task; DRT) and electroencephalographic (EEG; parietal alpha and frontal theta power, and the P3 event-related potential) measures for one group (n = 1 amputee participant, n = 10 non-amputee participants), and electrocardiographic (ECG; low/high frequency heart-rate variability ratio) and pupillometric (task-evoked pupillary response) measures for another group (n = 1 amputee participant, n = 10 non-amputee participants), because all measures could not reasonably be recorded simultaneously. Participants of both groups also completed the subjective NASA Task-Load Index (TLX) survey.

Ease of use, setup, piloting, and analysis complexity varied among measures. The DRT required minimal piloting, was simple to set up, and used straightforward analyses. ECG measures required moderate piloting, were simple to set up, and had somewhat complex analyses. Pupillometric measures required extensive piloting but were simple to set up and relatively simple to analyze. EEG measures required extensive piloting, extensive setup and equipment, careful monitoring, and moderately complex analyses.

Across subjects, the DRT, low/high frequency heart-rate variability ratio, task-evoked pupillary response, and NASA TLX significantly differentiated between the easy and hard tasks, whereas EEG measures (alpha power, theta power, and P3 event-related potential) did not. Aside from the NASA TLX, the DRT was the easiest to use and most sensitive to cognitive load across and within subjects. Among physiological measures, we recommend ECG, pupillometry, and EEG/ERPs, in that order.

This study provides the first evaluation of multiple objective and quantified cognitive workload measures during the same task with prosthesis use. User-focused cognitive workload assessments may increase our understanding of human interactions with advanced upper-limb neuroprostheses and facilitate their improvements and translation to real-world use.

**Significance Statement:** The human arm is dexterous and able to sense objects it contacts. Restoring sensory and motor function to a person with limb loss presents multiple challenges and requires improvements in robotics, biological interfaces, decoding biological signals for prosthesis movement, and sensory restoration. The scientific and engineering communities have made progress toward restoring arm function through advanced neuroprostheses. However, most studies focus solely on task performance, and they typically employ artificial experimental paradigms in which the user can devote full attention to the task, which is often unrealistic for use in everyday activities. To develop neuroprostheses capable of restoring intuitive arm function, engineers and scientists must also consider the difficulty of use, or cognitive burden, of using the neuroprosthesis. Although many measures of cognitive workload have been developed, few studies directly interrogate cognitive workload during neuroprosthesis use. An engineer or scientist seeking to employ cognitive workload measures during neuroprosthesis use will likely wonder, as we did, which measures are most suitable for their needs. To address this question, we empirically assess the practical and functional merits and limitations of several physiological, behavioral, and subjective techniques to measure cognitive workload during use of an advanced prosthesis. We anticipate that these findings may influence other medical and consumer areas of human-computer interaction, such as virtual reality or exoskeleton use.

## Introduction

Upper-limb prostheses generally rely on unintuitive controllers and do not restore sensation to the user, often resulting in prosthesis abandonment (Biddiss and Chau, 2007a). These limitations are among the major factors (Biddiss et al., 2007; Espinosa and Nathan-Roberts, 2019) in the high (30% to 50%) prosthesis abandonment rate (Pons et al., 2005; Biddiss and Chau, 2007b). More recent innovations, such as advanced, multi-articulating prostheses, have not yet produced substantial reductions in prosthesis abandonment (Salminger et al., 2020). Sophisticated solutions for restoring sensation (Tan et al., 2014; Graczyk et al., 2018; D’Anna et al., 2019; George et al., 2019; Schofield et al., 2019; Mastinu et al., 2020), improving motor control (Ortiz-Catalan et al., 2014b; Hargrove et al., 2017; Ameri et al., 2019; Salminger et al., 2019; Vu et al., 2020; Paskett et al., 2021), and improving prosthesis comfort through interventions such as osseointegration (Ortiz-Catalan et al., 2014a; Mastinu et al., 2020) provide valuable steps toward increasing user satisfaction and reducing abandonment.

One aspect of upper-limb prosthesis improvements that rarely is directly studied is the cognitive workload or effort required to use a prosthesis. Previous work (Resnik et al., 2012) has conveyed the need for direct cognitive workload measures for prosthesis use. Most studies with advanced prostheses demonstrate some form of performance improvement; however, performance does not necessarily imply ease-of-use and desirability. Ultimately, translating neuroprostheses from the laboratory to the clinic for long-term use will require the technologies to be *desirable*. Desirability will very likely increase with higher performance systems; it will certainly increase with high-performance systems that are easy to use. We found strong subjective preferences for certain movement decoders even though the objective performance was similar (Paskett et al., 2021), implying that user satisfaction and the desirability of the decoder was influenced by more than performance alone. Humans move their endogenous hand with dexterity and very little cognitive effort. That is, most movements are executed with a great deal of automaticity, without occupying the mind with the low-level details of the action. The ideal prosthesis should restore such automaticity to the user, enabling them to extend their focus beyond the prosthesis when carrying out a task. Quantifying cognitive workload during prosthesis use may provide a clearer path toward restoring automaticity.

Interrogating cognitive workload is possible through subjective, behavioral, and physiological measures. There are benefits and limitations to each. In the upper-limb prosthesis domain, most attempts at measuring cognitive workload (Markovic et al., 2018, 2020; Thomas et al., 2019) have been through subjective measures, such as the NASA TLX survey (Hart and Staveland, 1988). Subjective measures are quick and simple; however, they can suffer from large inter-individual variability, recall bias (Zahabi et al., 2019), and task-order dependency (McKendricka and Cherry, 2018). A few studies have employed behavioral measures (Witteveen et al., 2012; Raveh et al., 2018b; Valle et al., 2020) that generally use the performance in a secondary task (e.g., a memory task) as an index of difficulty of the primary (prosthesis) task. Behavioral measures are appealing because they measure cognitive workload contemporaneous with the prosthesis task and do not suffer as directly from recall bias or task-order dependency. However, they make the assumption that the paired secondary task will use mental capacity spared by the primary task and that trade-off strategies are not employed during the tasks (Fisk et al., 1983). Some studies have used physiological measures (Gonzalez et al., 2012; Deeny et al., 2014; White et al., 2017; Parr et al., 2019; Thomas et al., 2021) to quantify cognitive workload. Physiological measures are valuable because they rely on subconscious mechanisms to quantify cognitive workload and are relatively unaffected by experimenters’ or subjects’ biases or expectations. However, capturing these phenomena generally requires sophisticated equipment and well-prepared, and ofttimes constrained, conditions.

The question therefore arises: Which approach(es) should one use to measure cognitive workload? To answer this question, we used an ordinary prosthesis control task – matching a virtual hand to a target on a screen – for which we could easily manipulate task difficulty in order to compare subjective, behavioral, and physiological measures of cognitive workload. By collecting multiple cognitive workload measures during the same prosthesis task, our results facilitate direct comparisons of the measures’ effectiveness and utility. The results presented herein may thus aid researchers in selecting quantified cognitive workload measures for their own studies with advanced prostheses. Additionally, they may facilitate development, implementation, and clinical translation of easy-to-use prostheses.

## Methods

### Participant Recruitment

The present study was completed with two groups. In one group, we recorded behavioral and EEG measures of cognitive workload. One amputee participant, male, in his 40s, had a congenital left amputation approximately 10 cm below the elbow. Ten non-amputee participants completed the study: three female, seven male, 24.6 ± 3.4 years old, one left-handed, nine right-handed.

In the other group, we recorded cardiac and pupillometric measures of cognitive workload. One amputee participant, male, in his 40s, had bilateral traumatic amputations 10 years prior, about 8 cm below the elbow, and is right hand dominant. Ten non-amputee participants completed the study: five female, five male, 27.3 ± 12.2 years old, all right-handed. No participant from the first group was included in the second group, so that both groups had equally naïve participants.

### Cognitive Workload Measure Overview

We first briefly introduce the measures we employed in this study. For more in-depth reviews, we recommend (Charles and Nixon, 2019; Lohani et al., 2019).

#### DRT

The DRT is a secondary task in which a visual, auditory, or tactile stimulus prompts the user to respond by pressing a button. As the primary task increases in difficulty, the response time typically increases, and stimulus detection rate typically decreases (i.e., the user does not respond). An ISO standard of the DRT (ISO 17488:2016, 2016) has been used extensively in driving contexts (Ranney, T. A., Baldwin, G. H. S., Smith, L. A., Mazzae, E. N., & Pierce, 2014; Chang et al., 2017; Stojmenova et al., 2017; Strayer et al., 2017; Stojmenova and Sodnik, 2018), in which the stimulus is presented at random intervals of 3-5 s. More broadly, secondary tasks have been applied to prosthesis tasks with promising results, such as auditory discrimination tasks (Witteveen et al., 2012), memory tasks (Valle et al., 2020), and games (Raveh et al., 2018b, 2018a). In contrast with the referenced uses of secondary tasks, we find the DRT attractive because trials are collected rapidly (every few seconds) and the response times are nearly continuous. The DRT has not been applied previously to prosthesis research.

#### EEG and Event-Related Potentials

EEG is the measure of electrical potentials produced by the brain at the scalp surface. Alpha waves (8-12 Hz) in parietal regions indicate cortical idling and alpha power decreases with increased task demands (Keil et al., 2006). Theta waves (4 - 7 Hz) in frontal midline regions arise when cognitive control is required for a task (i.e., the task cannot be completed through an automatic strategy). Two studies have measured alpha waves (Gonzalez et al., 2012; Parr et al., 2019), but no study has analyzed theta waves during prosthesis use.

Event-related potentials (ERPs) are the brain’s electrophysiological response to a particular sensory, cognitive, or motor event (Luck, 2005). ERPs contain several components that represent various stages in neural processing of an event. When used to measure cognitive workload, ERPs are usually elicited through a secondary task, such as a DRT (Strayer et al., 2014). The P3 component, a positive potential arising roughly 300-ms post-stimulus, decreases in amplitude as the primary (in our case, prosthesis) task increases in difficulty and requires more resource allocation (Luck, 2005). Only one previous study has used ERPs as a cognitive workload measure during prosthesis use (Deeny et al., 2014).

#### Pupillometry

The eyes have been described as the “visible part of the brain” (Hess and Janisse, 1978). Pupillometry is the continuous measure of pupil size over the course of a task. Pupil size increases with cognitive demands, demonstrated as early as the 1960s (Kahneman and Beatty, 1966). Because pupil size changes due to several environmental, neurological, and psychological factors, trial averaging is often used to produce a “task-evoked pupillary response” (Beatty, 1982). Measuring the percentage of pupil dilation provides a measure that is robust to inter-individual and inter-trial baseline pupil size differences (Payne et al., 1968). Pupillometry has been used only rarely in the prosthesis domain (White et al., 2017; Zahabi et al., 2019).

#### Electrocardiography

Electrocardiography (ECG) is the measure of electrical potentials produced from the heart. Several time-domain and frequency-domain measures are sensitive to cognitive workload (Charles and Nixon, 2019). For our study, we used the low frequency (0.02 – 0.06 Hz) to high frequency (0.15 – 0.5 Hz) ratio (LF/HF ratio) because it showed the greatest sensitivity in our pilot experiments. One other prosthesis study has used ECG to measure cognitive workload (Gonzalez et al., 2012).

#### NASA TLX Survey

The NASA TLX is a subjective survey designed to measure perceived workload (Hart and Staveland, 1988) through six different categories on a 100-point scale: mental demand, physical demand, temporal demand, performance, effort, and frustration. Participants compare categories pairwise based on perceived importance in the task, and individual weightings from these comparisons are used to produce a composite score. The TLX has been used widely across many domains, including prostheses (Gonzalez et al., 2012; Markovic et al., 2018, 2020; Shaw et al., 2019; Thomas et al., 2019).

### Experiment Overview

Participants controlled a virtual prosthetic hand using sEMG signals to complete a virtual target task at easy and hard difficulties (Fig. 1). During the virtual task, we recorded subjective (NASA TLX), physiological (ECG, EEG, and pupillometry) and behavioral (DRT) data to be used as measures of cognitive workload. Because all the measures could not reasonably be collected at the same time, we recorded ECG and pupillometry together in one set of experiments, and EEG and the DRT together in another set of experiments.

**Figure 1.**
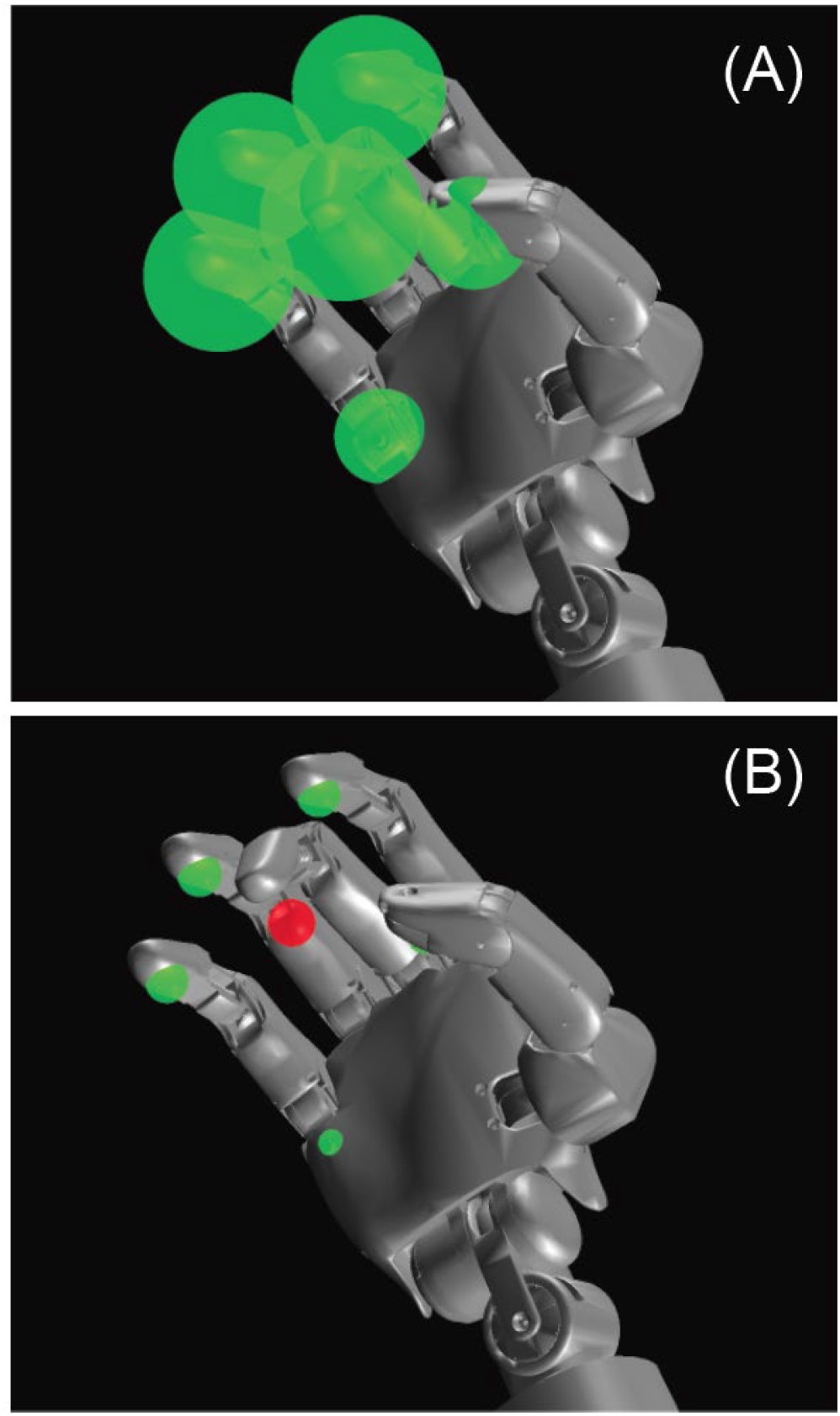
Virtual Target Task. Participants control the virtual hand and attempt to keep all targets green as random targets move to a target position for a specific time (5-15 s). (A) Large target with the middle finger active. Because the middle finger is within the target window, the target is green. (B) Small target with the middle finger active. Because the finger is outside the target window, the target is red.

#### Prosthesis Control

The prosthesis control methodology used in this study has been described previously (George et al., 2020a). In brief, sEMG was collected from an sEMG sleeve (George et al., 2020b) with the Grapevine System (Ripple Neuro LLC, Salt Lake City, UT). Thirty-two single-ended channels were acquired at 1 kHz and band-pass filtered between 15 Hz and 375 Hz with 4^th^-order Butterworth filters, and 60, 120, and 180 Hz 2^nd^-order Butterworth notch filters. After the sEMG sleeve was connected to the acquisition device, channels were manually inspected and removed if broken channels were detected (generally less than two channels). The differential pairs of all monopolar channels were calculated, and features (single-ended and differential) were created at 30 Hz using the mean absolute value of a 300-ms buffer (i.e., 528 features from an overlapping 300-ms boxcar filter). At 30 Hz, the buffer is updated every 33 ms. This update rate and buffer length has been used by our group extensively (George et al., 2018, 2020a).

sEMG was collected as participants mimicked preprogrammed movements of the virtual MSMS hand (Davoodi and Loeb, 2011). The preprogrammed movements consisted of index, middle, and ring finger flexions. Each movement consisted of a 0.7-s transition to flexed position, 4-s hold, and 0.7-s return to rest position. Participants completed two trials of each flexion as practice to gain familiarity with the virtual environment. After familiarization, participants completed five trials of each movement. Using a Gram-Schmidt forward selection algorithm (Nieveen et al., 2017), 48 sEMG features were selected as inputs to the decoder, a modified Kalman filter (George et al., 2020a). The 48 features and virtual hand kinematics were used to fit the parameters of the modified Kalman filter. After fitting the modified Kalman filter, users were given control over the virtual hand. We let the participants spend a few minutes exploring the control; in cases where the participants struggled to fully flex the fingers, participants repeated the five-trial mimicry and the modified Kalman filter was refitted.

#### DRT

We made a custom DRT system that interfaced with the Ripple Grapevine Digital I/O board. This system turned the tactile buzzer, a 10 mm x 2 mm vibration motor on a 4.5 V power supply, on or off when an output of the Digital I/O was set to high or low, respectively. The response button, when depressed, was recorded by the Ripple Grapevine system. The DRT vibrations were set to 1 s and turned off if the user pressed the response button before the 1 s had ended. Timestamps for the Digital I/O board are recorded at 30-kHz resolution. The DRT system was placed on the table near the participant and two separate cables for the vibration motor and response button were routed to the participant. We attached the DRT vibration motor to the collarbone with medical tape, opposite the hand used for the prosthesis task. We attached the response button to the index finger using a hook and loop fastener.

#### EEG & ERP Recordings

EEG was recorded based on the standard 10-20 system using a 34-electrode cap (Ripple Neuro LLC, Salt Lake City, UT). Electrode locations were: FP1, FP2, F7, F3, Fz, F4, F8, AFz, FT7, FT8, FC3, FCz, FC4, T3, C3, Cz, C4, T4, CP3, CPz, CP4, T5, P3, Pz, P4, T6, O1, Oz, O2, A1, A2, VEOL, HEOR, HEOL. The online reference was on electrode CPz, and the ground was AFz. We used Electro-Gel™ to bridge the connection between the electrodes and the scalp. Impedances of the electrodes were brought below 10 kOhm, typically close to 5 kOhm, using gentle scalp abrasion. We recorded the scalp EEG at 1 kHz and band-pass filtered between 1 Hz and 125 Hz with 4^th^-order Butterworth filters.

#### Pupillometry Recordings

Pupil diameter was recorded using the Pupil Labs’ Pupil Core head-mounted pupillometry device. We recorded pupil diameter with both pupil cameras at 120 Hz and 400×400 resolution. The 2D diameter output from the Pupil Labs’ software was used for analysis, which contains a measured diameter and the measurement confidence, ranging from zero to one. Room lighting was kept constant at approximately 100 lux, as measured by an Urceri MT-912 light meter.

#### ECG Recordings

ECG was recorded with the five-wire, four-lead Shimmer3 ECG unit. The unit recorded at 512 Hz. The Vx electrode was placed at V5, as suggested in the Shimmer3 ECG user manual, and the remaining electrodes were placed on the chest in the direction of the right arm, left arm, right leg, and left leg. ECG recordings were programmatically started and stopped when a target set was started or finished, respectively. The Shimmer3 logs the data onto an internal SD card, which was later extracted using Shimmer3 Consensys software.

#### Virtual Target Task

Participants completed a virtual target task in the MSMS virtual environment (Davoodi and Loeb, 2011) for easy and hard difficulties. In the virtual target task, a spherical target indicates the desired position of each degree-of-freedom. When a degree-of-freedom is within a specified radius of the target, the target is green; outside the allowable radius, the target is red. For our target tasks, the target was placed halfway through the movement window, with a target size (i.e., allowable radius) of 35% and 15% of the movement window for the easy and hard difficulties, respectively.

Participants were instructed to focus most of their attention on the active target, which was only one degree-of-freedom at a time. Participants were instructed that their objective was to keep the target green, not to keep the active degree-of-freedom in the middle of the target. Participants were encouraged to stay focused on the task and to avoid talking during the task in order to reduce cognitive demands beyond the task itself.

The subsequent sections describe the target task paradigm for each group. Because the different cognitive load measures have differing recording requirements, the experimental paradigms were slightly different for the two groups. The difficulty of the task (i.e., target size) was identical for both groups.

##### EEG & DRT Experiments

In the EEG and DRT virtual target task, the targets were active for 15 s. The participants first completed one practice set without the DRT that included one trial of each degree-of-freedom for each target size in a random order, for a total of six trials. For the next practice set, vibrotactile stimuli from the DRT system were presented randomly 3-5 s apart (uniformly distributed), according to ISO 17488 (ISO 17488:2016, 2016), resulting in, on average, 3 vibrotactile stimuli per active target. After the two practice sets, the participants completed eight rounds of the target task with the DRT. After the final target set was completed, users completed the NASA TLX for each target size in a random order.

##### ECG & Pupillometry Experiments

In the ECG and pupillometry virtual target task, the targets were active for 5 s with a random 3-5 s interval between targets (uniformly distributed). The participants first completed one practice target set for each target size. In the practice sets, each degree-of-freedom was tested twice, in a random order. After practicing the task, participants moved onto the full-length target sets. In one target set, each degree-of-freedom (index, middle, and ring finger) was tested six times, in a random order, for a total of 18 target trials per set. A set included only one target size. To calculate difference waves with the pupillary responses, participants also completed a “mimicry” set of targets. In the “mimicry” set, the computer perfectly completed the target task while the participants watched and mimicked the movements. Before the “mimicry” target set, participants were informed that the computer would be in control of the virtual hand, and they were instructed to watch the task and mimic the computer’s movements. The difference waves are discussed in greater detail in the analysis section. Participants completed one target set and one “mimicry” set for a single target size, then completed one target set and one “mimicry” set for the other target size. The initial target size was randomized. For the full experiment, participants completed four active and four “mimicry” target sets for each target size, resulting in 72 individual target trials per participant. After the final target set was completed for each target size, users completed the NASA TLX survey.

### Analysis

#### DRT

We analyzed the DRT according to the ISO standard (ISO 17488:2016, 2016). Responses (button presses after vibrotactile stimuli) less than 100 ms or greater than 2500 ms were counted as a miss. Response times more than three scaled median absolute deviations from the median were excluded from the analysis. We measured the hit rates and response times during each target size.

#### EEG & ERP

EEG was analyzed using EEGLAB v2021.0. The data were first resampled to 250 Hz. We re-referenced the electrodes to electrodes A1 and A2. The data were filtered from 0.1 Hz to 30 Hz using a second-order Butterworth filter. Artificial blink and horizontal eye movement channels were created by subtracting VEOL from FP1, and HEOL from HEOR, respectively.

For the frequency analysis, 15-s bins were created for the duration of the active target and separated by target size. Artifacts were detected and removed if the blink or horizontal movement channels exceeded a 100-µV threshold within a 200-ms sliding window. The 200-ms window passed across the 15-s bin in 50 ms increments. Individual bins were Hann-windowed prior to calculating the power-spectral density of each trial to avoid edge effects. Power-spectral densities of each trial were averaged together. The power for the alpha band (8-12 Hz) on electrode Pz and theta (4-7 Hz) band on electrode Fz were calculated. The percentages of power in the alpha and theta bands were calculated by dividing the power in the selected bands by the total power.

For the ERP analysis, bins were created from 200 ms prior to the buzzer onset to 1100 ms after the onset and separated by target size. Artifacts were detected and removed if the blink or horizontal movement channels exceeded a roughly 60-µV threshold within a 200-ms sliding window. The threshold was slightly adjusted when blinks or horizontal eye movements were not detected by the initial 60-µV threshold. The 200-ms window passed across the 1300 ms bin in 50 ms increments. Non-artifact trials were averaged together to produce an averaged ERP for each participant. The signed area was calculated from 200 ms to 650 ms (Strayer et al., 2014) to calculate the P3 ERP size. Averaged ERPs for each participant were averaged across participants to produce grand-averaged ERPs.

#### Pupillometry

Pupil recordings were aggregated by target size. We removed outliers defined as measurements greater than three scaled mean absolute deviations from the median of 60 samples (a 0.5-s window). We removed measurements with measurement confidence less than 0.8. Removed measurements were replaced with linearly interpolated values. Target trials with more than 20% low confidence measurements were removed from the aggregated set. The pre-trial baseline diameter, 1 s before the target became active, was subtracted from each trial. The percentage change in pupil size was calculated by dividing the response by the average size of the pupil during the 1-s pre-trial baseline. The baseline-subtracted pupillary responses of both eyes were combined and averaged to find the average pupillary response to the target task. The averaged pupillary response from the mimicry target (where the computer controlled the virtual hand) was subtracted from the averaged pupillary response to the active target (where the user controlled the virtual hand) to create a difference wave that would mitigate target-size dependent luminance effects in the response. We calculated the average value of the difference wave during the 5 s the target was active.

#### ECG

We obtained the LF/HF heart-rate variability ratio using the standard settings of PhysioZoo version 1.2.0 (Behar et al., 2018). The ECG was band-pass filtered from 3 Hz to 100 Hz with second and fifth-order Butterworth filters, respectively. Peaks in the ECG were detected using an energy-based QRS detector (Behar et al., 2014). The heart rate variability (intervals between normal heart beats) was calculated after removing outliers in the R-R peak intervals. Outliers were defined as intervals above or below 20% of the average of the moving window, which was 21 intervals. Power spectral density of the heart rate variability was calculated with Welch’s method. Finally, the LF/HF ratio was calculated by dividing the power in the low frequency region (0.04 Hz to 0.15 Hz) by the power in the high frequency region (0.15 Hz to 0.4 Hz).

#### Target Task Performance

We calculated the average percentage of time spent within the target window for each target size for each participant.

#### Statistical Procedures: Across-Subject

We tested the paired values derived from each measure for normality using the Shapiro-Wilk test. If the paired values were normally distributed, we used a paired t-test to show differences between the responses to the large and small targets. If the paired values were nonparametric, we used Wilcoxon’s signed-rank test. Because only one amputee participant completed each experiment, we did not include amputee participant results in our across-subject statistical measures and instead overlay results from amputee participants with the results of non-amputee participants.

#### Statistical Procedures: Within-Subject

Different measures may work well for some individuals, but not others. Additionally, due to the costs associated with implanting neural and electromyographic interfaces, it is common for studies to be completed with only a few subjects. We therefore were interested in the within-subject reliability of the cognitive workload measures. We completed within-subject analyses for each subject for each measure as appropriate for the measure and experimental paradigm. For the DRT, we conducted a two-sample t-test for all the DRT trials in an experiment. For the EEG & ERP measures, we conducted paired-sample t-tests with the average response for each of the eight rounds of the target task. For the pupil & ECG measures, we conducted paired-sample t-tests with the average response for each of the four rounds of the target task. We calculated the p-value from the statistical test and the absolute effect size using Cohen’s D for each participant. We report the median and first and third quartiles of p and D across non-amputee participants and the individual outcomes for the amputee participant. We report the number of participants for whom p < 0.05 by measure.

## Results

In brief, several but not all measures of cognitive load differentiated between the easy and hard tasks reliably in the aggregate intact subject pool. Significant differences (p < 0.05 or less) occurred for DRT, pupil dilation, LF/HF ratio, and TLX scores. Averaged ERPs, alpha and theta EEG powers, task-evoked pupillary responses, and heart-rate variability powers for the easy and hard tasks are shown in Fig. 2. Outcomes from individual participants are shown in Fig. 3. Each measure is discussed in detail in the following subsections. Parametric statistics are reported as mean ± standard error of the mean, and nonparametric statistics are reported as median [inter-quartile range].

**Figure 2.**
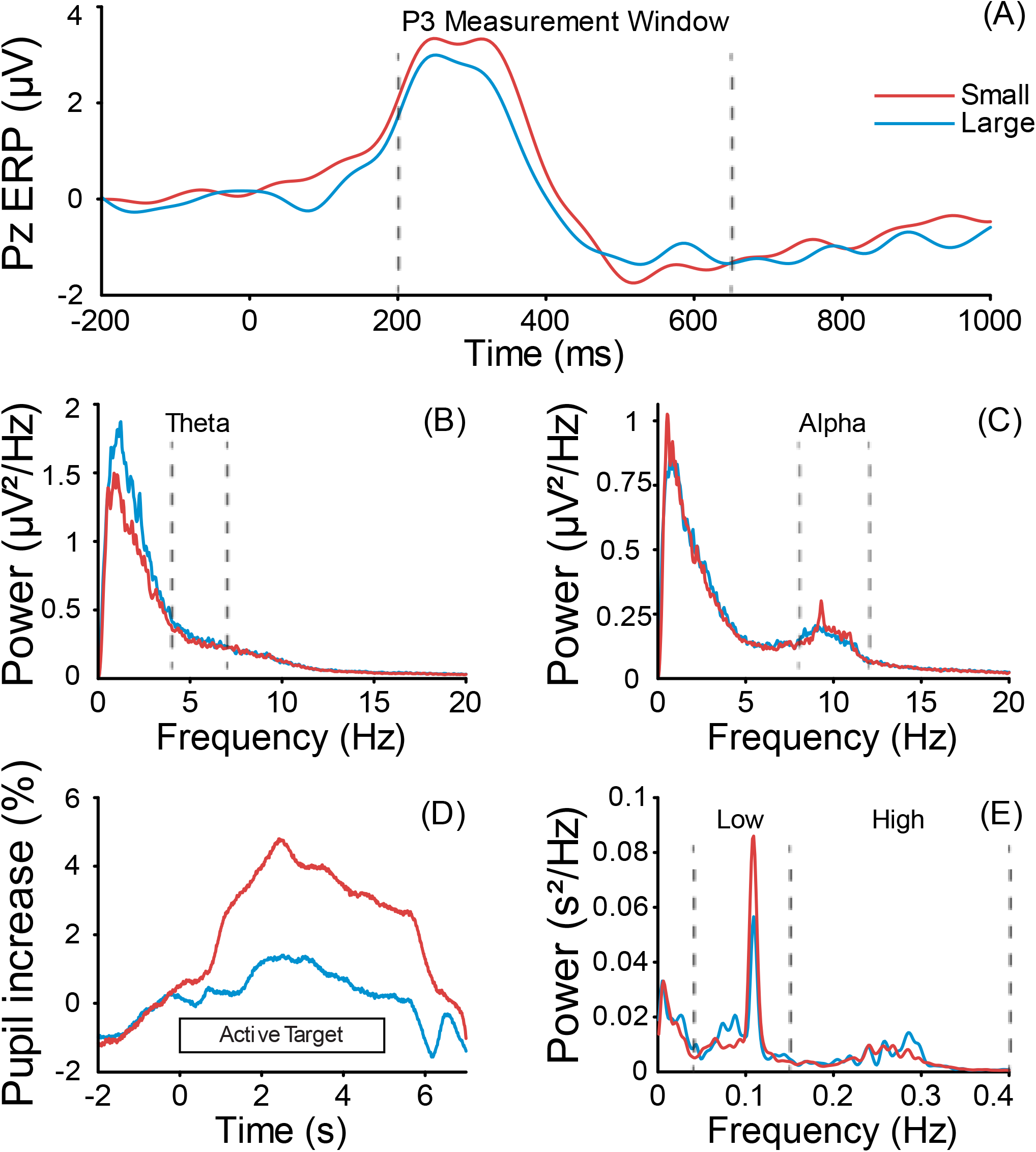
Raw physiological measures of cognitive load acquired during virtual target task at easy (large) and hard (small) difficulties for non-amputee participants. (A) Event-related potential (ERP) at electrode Pz arising from vibrotactile DRT stimulus. (B) Theta EEG power (4-7 Hz) at electrode Fz. (C) Alpha EEG power (8-12 Hz) at electrode Pz. (D) Luminance-corrected task-evoked pupil response. (E) Heart-rate variability power.

**Figure 3.**
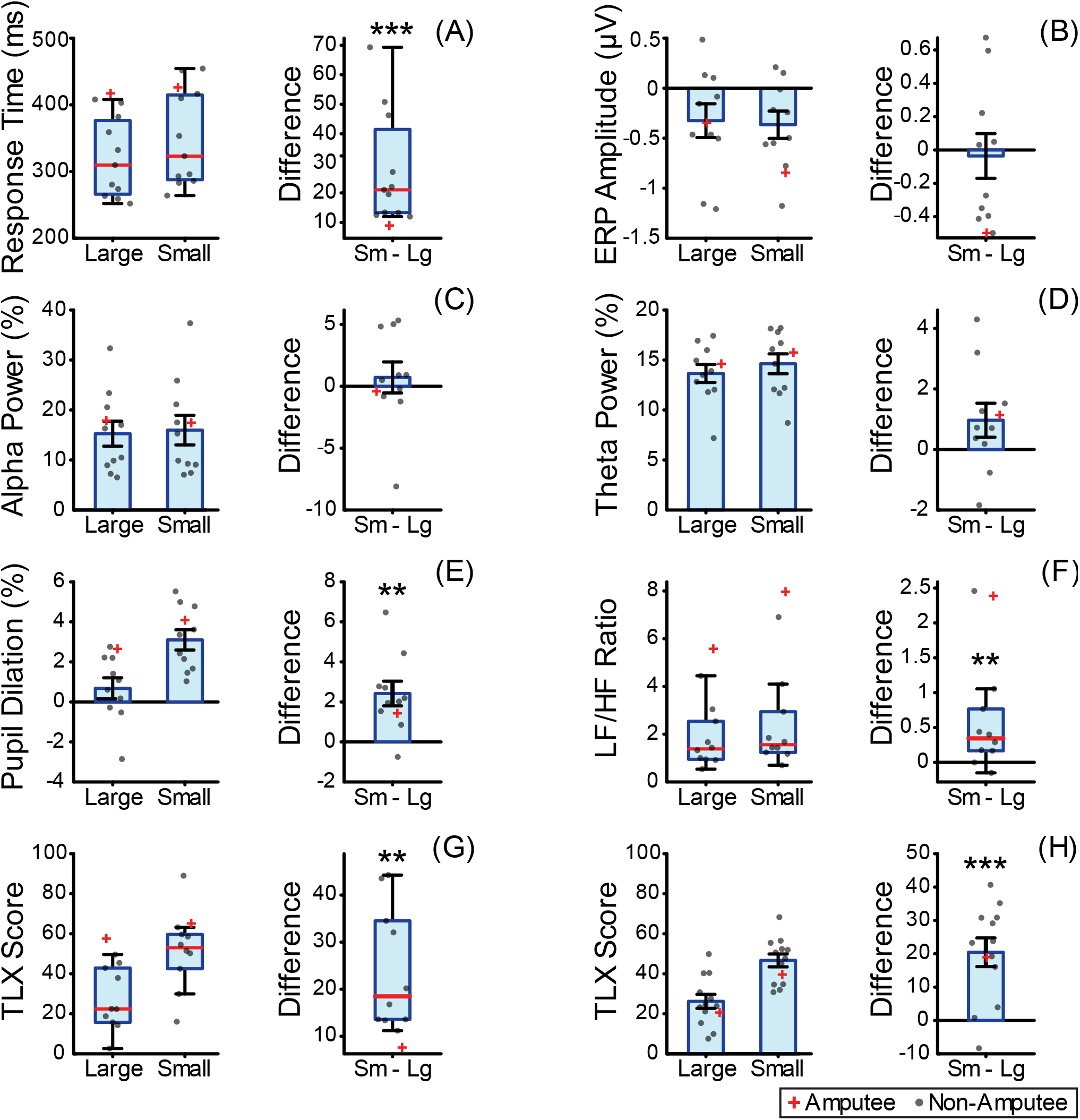
Several but not all measures of cognitive load changed with task difficulty. Shown are cognitive load measures from the (A) DRT, (B) P3 event-related potential, (C) alpha EEG power, (D) theta EEG power, (E) pupil dilation, (F) heart-rate variability low/high frequency ratio, and NASA TLX scores from the (G) EEG & DRT set and the (H) ECG & pupillometry set. Group descriptive and inferential statistics are depicted for the non-amputee participants only, without data from the amputee subject. For boxplots, red lines represent the median, the box represents Q1 and Q3, and the whiskers represent the outermost non-outlying points, as defined by the 1.5 * interquartile range extending from Q1 and Q3. For bar graphs, the top of the bar represents the mean, and the error bars represent the standard error of the mean. Paired comparisons were made (right subfigures) using parametric or nonparametric statistical tests, as applicable, for non-amputee participants only. *, **, and *** represent p < 0.05, p < 0.01, p < 0.001, respectively.

### Target Task

Confirming empirical differences in task difficulty, non-amputee participants performed significantly worse on the hard task (i.e., small target) compared with the easy task. For the DRT and EEG paradigm, non-amputee participants spent, on average, 33% ± 3% less time within the target window on the hard task (p < 0.001, paired t-test). The amputee participant had similar performance to the non-amputee participants, spending 41% less time within the target window for the hard task (small target: 48% [33%]; large target: 89% [9%]; p < 0.001, Wilcoxon’s rank sum test).

For the ECG & pupillometry paradigm, non-amputee participants spent 36% ± 2% less time within the target window for the hard task (p < 0.001, paired t-test). The amputee participant had similar performance to the non-amputee participants, spending 33% less time within the target window for the hard task (small target: 47% [21%]; large target: 80% [9%]; p < 0.001, Wilcoxon’s rank sum test).

### DRT

Non-amputee participants’ response times to the vibrotactile stimulus significantly increased by 21 ms [28 ms] when participants were completing the hard task (p < 0.001, Wilcoxon’s signed-rank test; Fig. 3a). The amputee participant’s response times increased by 9 ms for the hard task (small target: 419 ms [90 ms]; large target: 410 ms [87 ms]), but the difference was not significant (p = 0.42; Wilcoxon’s rank sum test). Hit rates (i.e., responses between 100 ms and 2500 ms) for both conditions were above 98% for all participants with no significant differences.

### EEG & ERP

EEG power spectra and grand-averaged ERPs for non-amputee participants are shown in Fig. 2a-c. No EEG or ERP measures were found to differ significantly between the easy and hard tasks (Fig. 3b-d). Theta power was not significantly different between easy and hard tasks for non-amputee participants (mean difference, hard task - easy task, 1.0% ± 0.6%; p = 0.12, paired t-test) or for the amputee participant (mean difference, hard task - easy task, 1.1% ± 0.4%; p = 0.22, paired t-test). Alpha power was not significantly changed for non-amputee participants (mean difference, hard task - easy task, 0.7% ± 1.3%; p = 0.58, paired t-test) or amputee participant (mean difference, hard task - easy task, 0.4% ± 0.2%; p = 0.70, paired t-test) for the amputee participant. The ERP size significantly decreased by 0.5 μV ± 0.1 μV for the hard task for the amputee participant (p < 0.001; paired t-test), but there was no significant difference for the non-amputee participants (mean difference, hard task - easy task, 0.0 μV ± 0.1 μV; p = 0.8, paired t-test).

### Pupillometry

The task-evoked pupillary responses for the non-amputee participants are shown in Fig. 2d. The task-evoked pupillary response significantly increased by 2.4% ± 0.6% for the hard task for non-amputee participants (p < 0.01, paired t-test; Fig. 3e). The amputee participant’s pupil response was not significantly different (mean difference, hard task - easy task, 1.4% ± 1.4%; paired t-test).

### ECG

The heart-rate variability power spectrum is shown in Fig. 2e. The LF/HF ratio significantly increased by a median value of 0.34 (p < 0.01, Wilcoxon’s signed-rank test; Fig. 3f) for non-amputee participants. The LF/HF ratio for the amputee participant did not significantly differ (mean difference, hard task - easy task, 2.4 ± 2.0%; p = 0.32, paired t-test).

### NASA TLX

For the DRT and EEG paradigm, the TLX score significantly increased by a median value of 18 for non-amputee participants (p < 0.01, Wilcoxon’s signed-rank test; Fig. 3g), and increased by 7 for the amputee participant. For the ECG & pupillometry paradigm, the TLX score significantly increased by an average value of 20 for non-amputee participants (p < 0.001, paired t-test; Fig. 3h), and increased by 19 for the amputee participant.

### Within-Subject Analysis

The p-values and effect sizes for the different cognitive load measures are shown in Table 1 for within-subject analyses for non-amputee participants and the amputee participant. Consistent with statistically significant results for across-subjects analyses, the DRT was the most reliable for the within-subject analysis, being significantly different for eight of ten non-amputee participants, with a median p-value of 0.001. Although EEG alpha power and theta power were not significantly different in the across-subjects analyses, these measures were significantly different for 5 and 3 individual non-amputee participants, respectively. Pupil dilation and LF/HF ration were both significant for the across-subjects analyses, but showed significant differences for only 2 and 1 individual non-amputee participants, respectively. The ERP was not significant for any individual non-amputee participant, consistent with the lack of significantly different results in across-subjects analyses. In contrast, for the amputee participant, the ERP was the only measure to be significantly different between the easy and hard tasks. The NASA TLX was administered only a single time for each participant, so inferential statistical analyses were not possible on a per-subject basis. However, all amputee and non-amputee participants rated the small target task as harder.

**Table 1.**
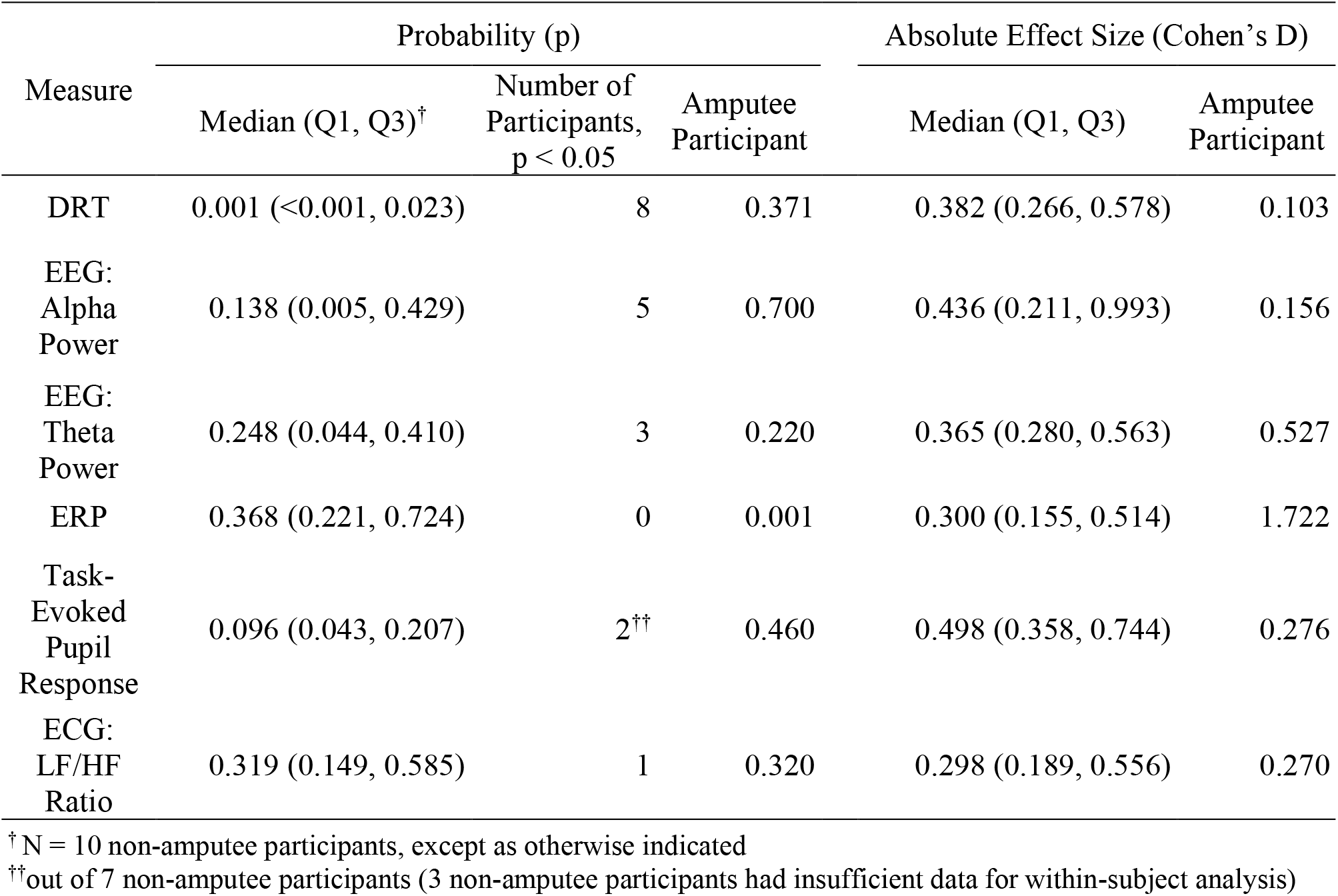
Within-subject reliability of the cognitive workload measures

| Measure | Probability (p) |  |  | Absolute Effect Size (Cohen's D) |  |
| --- | --- | --- | --- | --- | --- |
|  | Median (Q1, Q3)<sup>†</sup> | Number of Participants, p < 0.05 | Amputee Participant | Median (Q1, Q3) | Amputee Participant |
| DRT | 0.001 (<0.001, 0.023) | 8 | 0.371 | 0.382 (0.266, 0.578) | 0.103 |
| EEG: Alpha Power | 0.138 (0.005, 0.429) | 5 | 0.700 | 0.436 (0.211, 0.993) | 0.156 |
| EEG: Theta Power | 0.248 (0.044, 0.410) | 3 | 0.220 | 0.365 (0.280, 0.563) | 0.527 |
| ERP | 0.368 (0.221, 0.724) | 0 | 0.001 | 0.300 (0.155, 0.514) | 1.722 |
| Task-Evoked Pupil Response | 0.096 (0.043, 0.207) | 2<sup>††</sup> | 0.460 | 0.498 (0.358, 0.744) | 0.276 |
| ECG: LF/HF Ratio | 0.319 (0.149, 0.585) | 1 | 0.320 | 0.298 (0.189, 0.556) | 0.270 |

<sup>†</sup> N = 10 non-amputee participants, except as otherwise indicated
<sup>††</sup> out of 7 non-amputee participants (3 non-amputee participants had insufficient data for within-subject analysis)

## Discussion

This is the first prosthesis study that directly compares the efficacy and utility of several different objective, quantified cognitive workload measures that span physiological, behavioral, and subjective domains. Our objective was to determine the best technologies for user-focused prosthesis evaluations that will push laboratory developments toward clinical realities. We found the DRT to be the easiest to use and most sensitive to cognitive load across and within subjects. On the basis of their utility and their ability to differentiate among task difficulties, we next recommend ECG, pupillometry, and EEG/ERPs, in that order.

The comparative evaluations herein can inform the field’s use of cognitive workload measures in subsequent studies. Such studies could explore users’ responses to aspects of motor control, such as comparing decoders, finding a desirable number of degrees-of-freedom, or showing potential benefits of an active wrist. On the sensory side, one could explore the cognitive implications of sensorized and non-sensorized prostheses, compare feedback modalities (electrical vs. vibrotactile) or compare stimulation algorithms.

Designing experiments that could accommodate the recording requirements of the various measures used in this study was challenging because design choices could preferentially benefit a particular measure. We strived to provide suitable environments for all the measures and an experimental design that would enable effective collection of all the cognitive workload measures used. In the end, however, we were seeking for measures that are robust to environmental and experimental changes. We discuss the results, strengths, and limitations of the individual measures in the following subsections.

### DRT

We found that the DRT resulted in the most significant differentiation between the easy and hard tasks and was the most reliable for within-subject analysis. Overall, we recommend the DRT as a very reliable measure of cognitive workload that requires minimal setup and technical expertise. The DRT required minimal piloting and experimental manipulations before moving forward with recorded experiments. The DRT has several desirable characteristics as a cognitive workload measure: it is portable, requires minimal setup and its results are easily interpreted. This study demonstrates the first application of the DRT to a prosthesis task.

The DRT is limited by requiring physical button presses; however, many tasks for quantifying prosthesis performance are completed with one hand. Additionally, the response button could be modified for two-handed tasks (e.g., placed at the foot). The strengths and limitations of the DRT are discussed further in (Stojmenova and Sodnik, 2018). There are some aspects of behavioral measures that are not as attractive as physiological measures; however, the sensitivity and robustness of the DRT overcame our bias for physiological measures.

### ECG

The LF/HF ratio reliably detected differences in task difficulty. We recommend ECG, specifically the LF/HF ratio, as a viable physiological measure of cognitive workload that works for short-duration tasks. Although ECG worked well across subjects, for within-subject reliability, a greater number of trials is likely required. ECG is a relatively simple signal to obtain. The vast number of heart rate and heart-rate variability metrics (see (Charles and Nixon, 2019) for a review containing several ECG measures of cognitive workload) created a large parameter space to explore. Deciding on an ECG measure required a fair amount of piloting before use in experiments for the present study. Once selected, the LF/HF ratio remained robust. ECG measures of cognitive workload require relatively long recordings (>3-4 minutes), longer than many standardized prosthesis tasks, which can make task selection difficult.

In a study measuring cognitive load with a sensorized prosthesis, heart rate was found to decrease when participants had audiovisual feedback vs. visual feedback alone (Gonzalez et al., 2012). However, in the same study, heart-rate variability had no significant effect for any of the three conditions tested.

### Pupillometry

The task-evoked pupillary response successfully differentiated between the easy and hard tasks. With some reservation, we recommend pupillometry as a viable method of measuring cognitive workload during prosthesis use if the task can be modified for trial-averaged pupil responses. With no widely accepted continuous measure of cognitive workload, the task had to be time-locked to perform trial averaging. For the virtual target task, time-locking is straightforward; however, this is not the case with many physical prosthesis tasks. Additionally, pupillometry requires controlled luminance, which adds more complexity to experiment setup. Although pupillometry provided a robust response in the end, we had to pilot the experiments extensively and carefully design our analyses to uncover the effect.

Pupillometry actually resulted in the largest effect size on an individual basis, but was not as consistent across subjects. Setting up pupillometry was relatively simple; the head-mounted pupillometry system used was nonintrusive, and robust to head movements.

Two other studies have used pupillometry as a measure of cognitive workload during a prosthesis control comparison. One study showed that the number of pupillary increases was significantly different for direct and classifier prosthesis control (White et al., 2017). The other study showed that average pupil size was significantly different for a similar comparison (Zahabi et al., 2019).

### EEG & ERP

EEG and ERPs were lacking in sensitivity and diagnostic ability. Although we find the measures attractive, these barriers make it difficult to recommend using EEG & ERPs as reliable, easy-to-use measures of cognitive workload. Frontal theta power, a measure of cognitive control (i.e., when a task cannot be completed with an automatic, subconscious strategy) was close to a statistical trend across subjects, but parietal alpha power and the P3 ERP were far from any across-subject statistical trend. Alpha power was significantly different within-subject for five of ten participants, but the shift in power was inconsistent, resulting in no across-subject trend. The P3 response was surprisingly consistent for the amputee participant, highlighting the concept that different measures may work well for some persons but not others. EEG and ERPs are appealing as they are direct measures of neural activity; however, recording EEG and ERPs requires specialized training, time-consuming setup, and relatively expensive equipment.

In a similar virtual prosthesis task (Deeny et al., 2014), the P300 ERP differed between passively viewing the task and a hard condition, but there was no statistical difference between actively completing the task under easy or hard conditions. In a physical task evaluating a sensory feedback system, alpha power significantly differed between different feedback modalities (Gonzalez et al., 2012).

### NASA TLX

We found that the NASA TLX worked well with the target matching task, as can be reasonably expected when the task difficulty is quite obviously manipulated (i.e., it is very obvious to expect a small target to be more difficult than a large target). We recommend the TLX as a cognitive workload measure because of its simplicity, short duration, and widespread use. Because it is completed after a task is over and depends entirely on subjective self-report, the TLX suffers from recall bias (Zahabi et al., 2019), task-order dependency (McKendricka and Cherry, 2018) and other possible subjective biases. These effects can generally be mitigated through proper experimental design and participant instruction. However, some argue the TLX measures task difficulty more than it measures perceived mental workload (McKendricka and Cherry, 2018).

Many prosthesis studies have employed the NASA TLX for comparing movement decoders (Deeny et al., 2014; White et al., 2017; Osborn et al., 2021; Paskett et al., 2021) and sensory feedback (Gonzalez et al., 2012; Markovic et al., 2018, 2020; Thomas et al., 2021). The TLX generally provides a reliable response to changes in task difficulty.

### Conclusion

This study utilizes several physiological, behavioral, and subjective cognitive workload measures during a prosthesis task with known difficulty manipulations. Through collecting multiple measures during the same task, the study enables researchers to comparatively evaluate the effectiveness and utility of the various measures. Directly comparing several cognitive workload measures will aid neuroprosthesis researchers in applying cognitive workload to their own studies. Overall, we recommend the DRT, ECG, pupillometry, and EEG/ERPs, in that order, along with the traditional NASA TLX. EEG/ERP measures typically were not reliably informative across subjects, although some EEG measures worked will for a subset of individuals. Incorporating cognitive workload measures, and general user experience, to neuroprosthesis studies provides a path for better, more intuitive neuroprostheses which can more readily be translated to clinical realities.

## Data Availability

All data produced in the present study are available upon reasonable request to the authors

## Declarations

### Ethics approval and consent to participate

Participants completed the study after providing informed consent. All experiments were conducted under the oversight of the University of Utah IRB.

### Consent for publication

Not applicable.

### Availability of data and materials

The data that support the findings of this study are available from the corresponding author, MDP, upon reasonable request.

### Authors’ Contributions

MDP performed background research, developed the software, built the custom DRT, developed the experiments, conducted the experiments, analyzed the data, and wrote the manuscript. JKG performed background research, conducted the experiments, and analyzed the data. STJ performed background research, conducted the experiments, and analyzed the data. MRB developed the software, built the custom DRT, and helped revise the manuscript. TSD developed the majority of the software and helped revise the manuscript. CCD aided in participant recruitment, background research, and helped revise the manuscript. JMC aided in experiment development and provided cognitive load measurement expertise. DLS aided in experiment development and provided cognitive load measurement expertise. GAC oversaw all aspects of the study.

## Acknowledgments

We thank Dr. Brennan Payne, Dr. Trafton Drew, Sara LoTemplio, and Jack Silcox for their advice and expertise with developing this study.

We thank Ripple Neuro, LLC for providing EEG caps compatible with their neural interface processors, enabling the EEG recordings for this study.

